# Pointing in the right direction: Motor improvements with directional versus circular DBS

**DOI:** 10.1101/2023.02.27.23286503

**Authors:** Christopher P Hurt, Daniel J Kuhman, Barton L Guthrie, Joseph W Olson, Arie Nakhmani, Melissa Wade, Harrison C Walker

## Abstract

Directional brain stimulation provides greater control of radial current spread than conventional ring-shaped electrodes. Whether this added flexibility can improve motor function is unclear. Here we examine whether directional and circular stimulation differentially change motor performance in patients with Parkinson’s disease. We measured motor behaviors in 31 patients who underwent unilateral subthalamic nucleus brain stimulation surgery (SUNDIAL, NCT03353688) from each of 8 configurations (6 directional contacts and their corresponding rings) during device activation. Objective measures of dexterity, gait, balance, and overall mobility were queried in a double-blind fashion in the practically defined “off” medication state versus preoperative baseline with stimulus amplitude at the center of the therapeutic window. The best versus the worst directional contact on a given row yields significant changes in performance across 5 motor tasks (p<0.001 each task). Specific stimulation directions can worsen function versus baseline, whereas the best direction yields greater improvement than ring stimulation (p=0.005, p=0.001, p=0.007, p<0.001, respectively, across tasks). Although directional DBS improves side effect thresholds versus ring stimulation (p<0.001), the side effect threshold and therapeutic window correlate only modestly with motor improvements. Resting beta power did not predict motor improvements by directional DBS across any of the motor tasks. Optimized directional subthalamic nucleus DBS yields better group-level motor performance than ring stimulation, in addition to known advantages related to tolerability. Prospective studies should evaluate whether these improvements persist over longer time intervals.

## Introduction

Deep brain stimulation (DBS) is effective for motor symptoms of Parkinson’s disease (PD) in patients who no longer respond optimally to pharmacologic therapies.^1^ The goal of both surgical targeting and clinical programming is to maximize motor improvements and avoid or reduce potential stimulation side effects. However, while beneficial at the group level, conventional ring-shaped DBS leads yield a circular electrical field that can unintentionally interact with off-target structures. Recent advances in lead architecture allow greater control over stimulation in the axial dimension, leading to improvements in therapeutic window (the range of effective DBS amplitudes at a given stimulation site) and side effect thresholds.^2-4^ With this increased flexibility, directional leads substantially increase programming time and complexity, prompting efforts to identify biomarkers such as resting beta frequency to predict the contacts with greatest efficacy^5^. However, also important is the extent to which directional leads impact functional motor outcomes.

Motor improvements from DBS can vary widely from minimal^6-8^ to large and clinically significant.^9^ Response variability across individuals arises from differences in neuroanatomy, stimulation site, electrode architecture, disease phenotype, assessment methods, and other factors.^10,11^ In a prior study, we found that changes in DBS parameters elicit reliable changes in walking speed within participants ranging from meaningful improvements to worsening versus preoperative baseline.^9^ This suggests that walking speed and similar metrics from other functional domains might display similar variations in outcome to directional stimulation.

A prior study in ten participants with bilateral STN leads suggests that directional DBS improves Movement Disorders Society - Unified Parkinson Disease Rating Scale (MDS-UPDRS) items related to contralateral hand rotation but not rigidity or repetitive finger tapping.^4^ Other authors suggest minimal differences in motor improvement with directional versus ring stimulation with optimal lead placement.^3^ Despite its greater complexity, directional DBS is already adopted widely in many centers^12^, with approximately 2/3 of patients receiving directional stimulation rather than conventional ring stimulation in one series.^13^ Better understanding how directional stimulation impacts motor performance has important implications for both clinical programming and the design and parameterization of future devices. If directional stimulation provides better group-level motor performance than ring stimulation, this provides a stronger rationale for utilizing directionality from the outset, rather than merely as a ‘backup’ strategy.

Despite its immense overall value, individual UPDRS motor items are relatively coarse ordinal scores assigned by a subjective rater with outcomes that sometimes correlate only modestly with quality of life.^14,15^ Moderate changes in limb rigidity and bradykinesia can be difficult to capture, especially when measured repeatedly in a single session. Similarly, UPDRS items related to gait, mobility, and balance can be insensitive to important changes in function and addressing declines in these domains is a major unmet need for therapy.^16,17^ Here we systematically tested motor performance during initial DBS activation with a multidimensional battery of objective, validated tasks. We tested three interrelated hypotheses: (1) directional DBS yields significant within- and across-participant changes in motor function, (2) optimized directional stimulation can yield greater improvements than conventional ring stimulation, and (3) therapeutic window and/or resting beta frequency (13-30 Hz) power predict motor improvements from directional DBS.^5^

## Methods

### Enrollment, DBS surgery, and local field potential acquisition

With prior Institutional Review Board approval from the University of Alabama and Birmingham and written informed consent, we studied 31 PD patients at a preoperative screening visit and initial device activation after surgery as part of the SUNDIAL study (NCT03353688, IDE# G170063, Table 1). Patients were approached for participation after our multidisciplinary committee recommended unilateral STN DBS as part of routine care. Inclusion required ≥30% improvement in the MDS-UPDRS part III on dopaminergic medications versus the practically defined “off” state (≥12 hours off medications), Hoehn and Yahr score >1, and Dementia Rating Scale-2 score ≥130. Exclusion criteria included duration of disease <4 years, and no history of stroke or other significant neurological conditions. Full enrollment criteria are available at clinical.trials.gov. After screening, participants underwent unilateral DBS surgery to treat the most affected side of the body, implanting a Boston Scientific Cartesia^™^ ‘1-3-3-1’ lead. Targeting during surgery was informed by multi-pass microelectrode recordings, macrostimulation, intraoperative O-arm 2 imaging, and macrostimulation with the newly implanted DBS lead. For external stimulation, we used a Multichannel Systems STG4008 8-channel optically insulated stimulus generator. Immediately after implant, we recorded local field potentials from each of the 8 DBS contacts at rest in two non-consecutive time intervals of one minute each. Participants were instructed to move as little as possible, while remaining awake and alert with eyes open. We sampled the local field potentials at 25 kHz over the entire two-minute interval using a Brain Vision actiCHamp amplifier without filters. Primary analyses and visualizations displayed local field potentials in common average reference (each DBS contact minus the mean of all other DBS contacts). Monopolar montages (each contact referenced to DBS 1, DBS 8, or a scalp electrode) and split bipolar montage to contact 1 for ventral directional contacts and 8 to dorsal directional contacts yielded essentially identical results as common average reference. We visually excluded brief episodes of spontaneous or incidental movements based on *post hoc* analyses of EMG signals from contralateral orbicularis oris, bilateral flexor carpi ulnaris, and contralateral gastrocnemius muscles. Resting beta power was computed as the mean area under the wavelet scalogram in the beta frequency range (13-30 Hz) for each DBS contact. One participant had corrupted field potential signals and was excluded from the spectral analyses.

**Table 1.** Demographics.

|  | gender | diagnosis | target | implant hemisphere | duration (years) | LEDD (mg) | UPDRS part 3 total |  |  |
| --- | --- | --- | --- | --- | --- | --- | --- | --- | --- |
|  |  |  |  |  |  |  | off | on | change |
|  | female | pd | stn | right | 10 | 930 | 53 | 16 | 69.8% |
|  | female | pd | stn | left | 11 | 1600 | 44 | 14 | 68.2% |
|  | female | pd | stn | left | 7 | 1070 | 54 | 20 | 63.0% |
|  | female | pd | stn | right | 10 | 470 | 34 | 14 | 58.8% |
|  | female | pd | stn | right | 7 | 915 | 48 | 22 | 54.2% |
|  | female | pd | stn | left | 9 | 2250 | 52 | 25 | 51.9% |
|  | female | pd | stn | right | 9 | 780 | 77 | 39 | 49.4% |
|  | female | pd | stn | right | 5 | 832 | 17 | 9 | 47.1% |
|  | female | pd | stn | right | 12 | 600 | 41 | 22 | 46.3% |
|  | female | pd | stn | left | 5 | 0 | 38 | 25 | 34.2% |
|  | male | pd | stn | left | 5 | 1540 | 42 | 12 | 71.4% |
|  | male | pd | stn | left | 11 | 1680 | 54 | 20 | 63.0% |
|  | male | pd | stn | left | 7 | 950 | 73 | 29 | 60.3% |
|  | male | pd | stn | left | 19 | 1200 | 53 | 25 | 52.8% |
|  | male | pd | stn | left | 10 | 1000 | 55 | 27 | 50.9% |
|  | male | pd | stn | left | 11 | 1200 | 59 | 29 | 50.8% |
|  | male | pd | stn | right | 8 | 750 | 46 | 23 | 50.0% |
|  | male | pd | stn | left | 12 | 1650 | 80 | 41 | 48.8% |
|  | male | pd | stn | left | 6 | 550 | 33 | 17 | 48.5% |
|  | male | pd | stn | right | 8 | 600 | 61 | 33 | 45.9% |
|  | male | pd | stn | right | 7 | 460 | 33 | 18 | 45.5% |
|  | male | pd | stn | left | 9 | 1300 | 48 | 27 | 43.8% |
|  | male | pd | stn | right | 11 | 1260 | 53 | 30 | 43.4% |
|  | male | pd | stn | left | 5 | 785 | 45 | 26 | 42.2% |
|  | male | pd | stn | right | 6 | 1260 | 47 | 31 | 34.0% |
|  | male | pd | stn | right | 4 | 1050 | 53 | 35 | 34.0% |
|  | male | pd | stn | left | 4 | 730 | 56 | 37 | 33.9% |
|  | male | pd | stn | right | 12 | 1000 | 53 | 36 | 32.1% |
|  | male | pd | stn | left | 4 | 1064 | 47 | 32 | 31.9% |
|  | male | pd | stn | left | 8 | 799 | 42 | 29 | 31.0% |
|  | male | pd | stn | right | 4 | 0 | 34 |  |  |
| mean | — | — | — | — | 8.3 | 976.6 | 49.2 | 25.4 | 48.6% |
| sd | — | — | — | — | 3.3 | 476.3 | 13.1 | 8.3 | 11.6% |

### Motor battery

We measured motor function in a biomechanics laboratory at baseline with a battery of objective, validated tasks, including elements from the NIH Toolkit.^18^ All assessments were performed in the practically defined “off” state at a screening baseline visit and approximately 1 month after surgery on the day of initial device activation.^19^ Domains of function included upper and lower extremity dexterity, gait, balance, and overall mobility. The 9-hole pegboard test measures upper limb dexterity, incorporating repetitive, combined usage of fine motor object manipulation and wrist, elbow, and shoulder movements. This is a timed task where individuals grasp pegs from a cup one at a time with the limb contralateral to the intended hemisphere for DBS, insert the pegs into the 9-hole pegboard, and then extract and place each peg back into the cup. We also created and validated an analogous timed task to assess lower limb dexterity.^20^ Seated participants use the contralateral lower limb to touch the numbers 1 through 9 placed in a 3×3 grid on a force plate embedded in the floor. They begin the task with their foot on an external target just posterior to the grid and sequentially tap each number in an ‘S-shaped’ pattern on the grid, returning to the target between numbers. To assess gait, participants walked over a 5.5 m gait mat (Zeno, Protokinetics, Havertown, PA) with 2 m before and after the mat, allowing participants to reach and maintain comfortable walking speed. To assess overall mobility, we used the Timed Up and Go test. Participants stand from a seated position in a chair, walk 3 m, turn around, and return to a seated position, while avoiding using their hands to stand up or sit down. To assess static balance, we asked individuals to stand unshod on a force plate (AMTI, Watertown MA) with minimal voluntary movement while fixating on an ‘X’ on the wall while the center of pressure of the individual is quantified as an area measure in mm^2^.

### Monopolar review

Participants returned for initial device activation approximately 1 month after device implant. Participant and rater were blinded to stimulation parameters throughout, and sessions activating a single lead typically lasted 3 to 4 hours. An experienced DBS clinician programmed in monopolar cathodal configuration with pulse width 60 µs and frequency 130 Hz. The directional lead consists of 8 contacts arranged in a 1-3-3-1 pattern, with directional contact segments in the middle two rows. For our primary hypotheses, we assessed behavioral responses to DBS from the 6 directional contacts versus their corresponding virtual ring (i.e., activating the 3 directional contacts on a given row at 33% current fractionation each). Contacts were tested in a pre-specified, random order to mitigate potential order effects. We measured the therapeutic window at each DBS configuration.^21^ First we defined the therapeutic floor for a given stimulation site as the minimum stimulus amplitude providing significant symptomatic improvement on the contralateral body (e.g., relief of contralateral rigidity and/or rest tremor). DBS amplitude was gradually increased until a side effect emerged (typically contralateral muscle pulling, dysarthria, persistent or uncomfortable paresthesia, or worsening of parkinsonism). The therapeutic window ceiling is defined as 0.1 mA less than the amplitude threshold for this effect. Therapeutic window size is computed as ceiling minus floor (in mA). For motor testing at each stimulation site, DBS amplitude was set at the midpoint of the therapeutic window, or (ceiling + floor) / 2, for each contact. In instances where there was no therapeutic window, DBS amplitude was set to 0.2 mA beneath the threshold for side effects or 4 mA (whichever larger). We allowed a few minutes for acclimation to DBS at each site and then queried the motor battery. Between settings, we deactivated DBS and did not proceed with subsequent testing until visible return of motor symptoms (typically less than 5 minutes).

### Data analyses

Outcome variables of interest fall into three broad categories: performance on motor tasks, thresholds for stimulation effects (i.e., therapeutic window), and local field potential signals. Our primary hypothesis examines whether directional stimulation yields different motor performance versus conventional ring stimulation. We rank-ordered outcomes within a given row (i.e., dorsal and ventral) and quantified the best versus worst performance, also noting how frequently these changes met or exceeded the clinically important differences for each task (where known). Clinical studies have defined clinically important differences for the comfortable walk speed (0.06-0.22 m/s for a small to large effect),^22^ timed up and go (3.5 seconds),^23^ and 9-hole pegboard (2.6 seconds)^24^ tasks. To test our primary hypotheses on motor function, we assessed differences between directional and ring stimulation within a given DBS row using 2×2 repeated measures ANOVA. The dependent variable, motor performance, was expressed as a change versus preoperative baseline. For each ANOVA, if a significant difference was found with the omnibus test, we assessed pairwise effects with the Bonferroni method. We used identical statistical methods to evaluate how directional stimulation changes therapeutic window and side-effect threshold on a given DBS row. Finally, we also assessed the extent to which motor improvement is predicted by other motor tasks, stimulation thresholds, and resting beta power. We analyzed these relationships with the R package ‘rmcorr’ which measures the strength and significance of correlations among variables with repeated measures. Elements in the correlation matrix visualization from ‘corrplot’ are ordered mathematically in an unsupervised manner based upon the correlation structure of their angular eigenvectors with

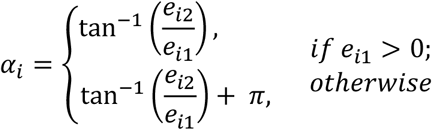

where *e*_1_ and *e*_2_ are the largest two eigenvalues of the correlation matrix. The significance threshold for all statistical tests was *p*<0.05.

## Results

### Motor performance with directional versus ring stimulation

Directional contacts significantly modify both upper and lower extremity dexterity in the 9-hole pegboard and lower extremity target tasks (Fig. 1, Fig. 5). Contrasting the best versus worst directional contact on a given DBS row for each participant, we measured a clinically significant change in performance in 80% (25/31) of dorsal (6.0 ± 4.3 sec) and 77% (24/31) of ventral contacts, (5.8 ± 4.3 seconds, Fig. 1, first column cyan lines). Contrasting directional versus ring contacts, the row by contact (ring versus directional) interaction and main effects for DBS row were non-significant (*p*=0.63 and *p*=0.37). Nine-hole pegboard performance was faster on the best directional contact versus its corresponding virtual ring on both the dorsal and ventral rows (*p*<0.01, respectively). For the lower extremity target task (Fig. 1), the best versus worst directional contact on a given DBS row yielded a difference of 2.5 ± 3.5 seconds (33% improvement) for the dorsal row and 2.6 ± 2.6 seconds (22% improvement) for the ventral row. The row by contact interaction and main effects of DBS row were non-significant (*p*=0.36 and *p*=0.87 respectively). Directional contacts versus virtual rings yielded better lower extremity dexterity at the group level on both the dorsal and ventral DBS rows, as well (*p*<0.01, respectively).

**Figure 1.**
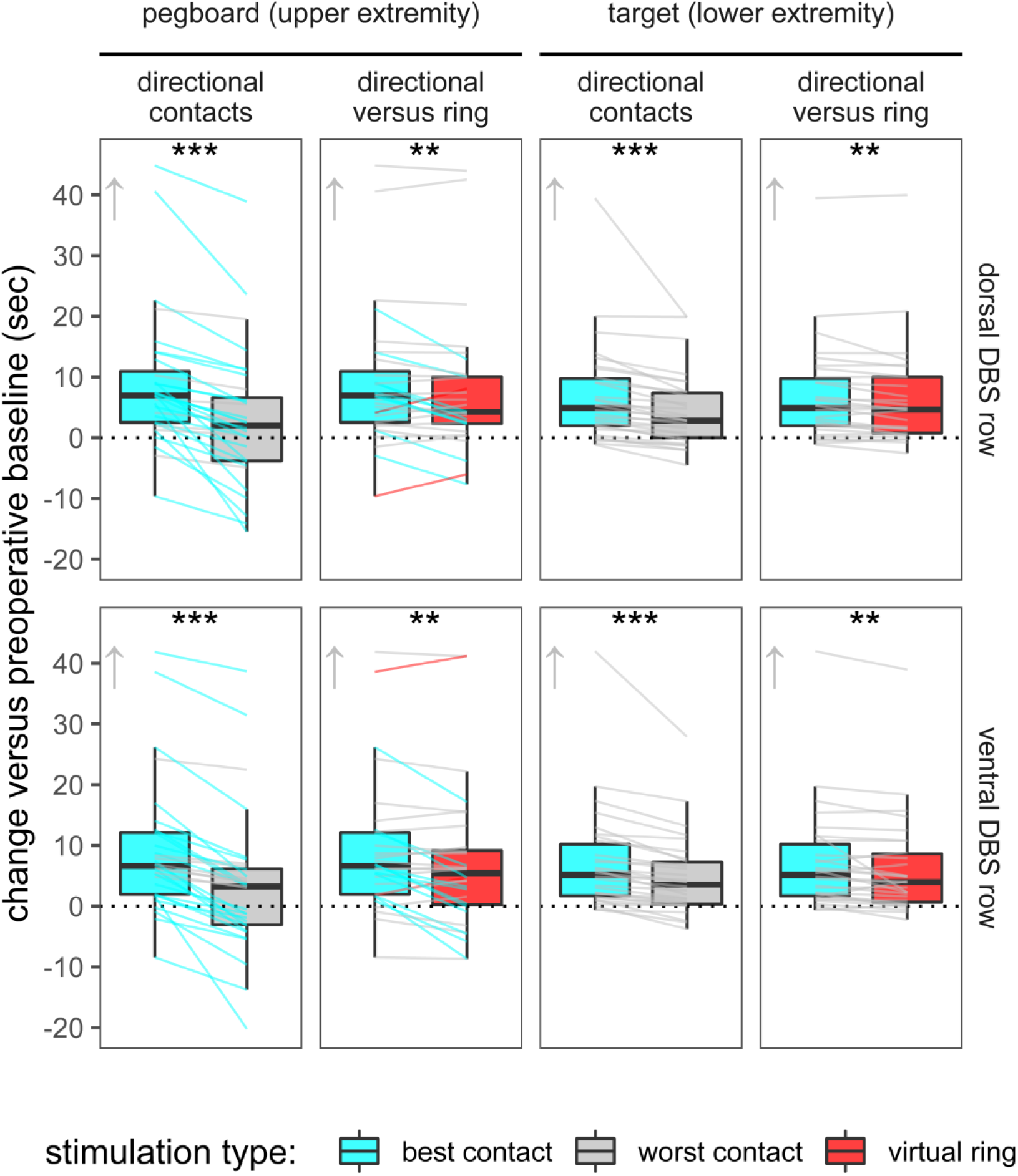
Dexterity tasks measure motor coordination and bradykinesia in the contralateral upper and lower limbs. Optimized directional parameters yield greater group-level improvement than ring stimulation versus baseline on both tasks (arrows indicate direction of improvement). Despite these improvements, directional and ring stimulation can sometimes worsen motor performance, and optimized directional DBS often rescues this worsening (line segments). Changes between directional and ring stimulation that meet or exceed the clinically important differences are highlighted with cyan and red lines, respectively. Clinical important differences is not established for the lower extremity task. * p <0.05, ** p<0.01, and *** p<0.001 for group-level contrasts.

Directional contacts significantly modify gait, overall mobility, and balance in the comfortable walk speed, timed up and go, and quiet stance tasks (Fig. 2., Fig. 5). Walk speed improved with the best versus worst directional contact on the dorsal and ventral DBS row in 61% (19/31) and 77% (24/31) of cases, respectively (Fig. 2 first column, cyan lines). Walk speed differed by 0.13 ± 0.14 and 0.11 ± 0.11 m/s on the best versus worst dorsal and ventral directional contacts, respectively. The row by contact interaction and main effects of DBS were non-significant (p=0.11 and *p*=0.16). However, we detected faster walk speeds during directional stimulation on both the dorsal and ventral rows (p<0.01 each, Fig. 2 second column). For timed up and go, we exceeded the threshold for minimal clinically significant improvement (≥3.5 seconds change) with the best versus worst directional contact in 13% (4/31) of both dorsal and ventral contacts (Fig. 1A first column, cyan lines). Mean differences between best and worst contacts were 2.0 ± 2.4 and 1.9 ± 1.7 seconds on the dorsal and ventral rows (Fig. 2). There was a non-significant row by contact interaction and main effect of DBS row (*p*=0.60 and *p*=0.98, respectively), but group level performance was faster for the best directional contact versus virtual ring on each DBS row (*p*<0.01 each). For the quiet stance task (Fig. 3, Fig. 5) sway area for the worst ventral contact and ring were each >5.5 standard deviations from the mean in one participant, thus these observations were omitted from group level analyses. Sway area differed by 403.7 ± 562.4 mm^2^ for best versus worst dorsal contacts (40% improvement) and 345.6 ± 439.0 mm^2^ for best versus worse ventral contacts (41% improvement). Row by contact interaction and main effect of DBS row were non-significant (*p*=0.47 and *p*=0.73 respectively) but best directional contact again outperformed virtual ring on each DBS row (*p*<0.01 each).

**Figure 2.**
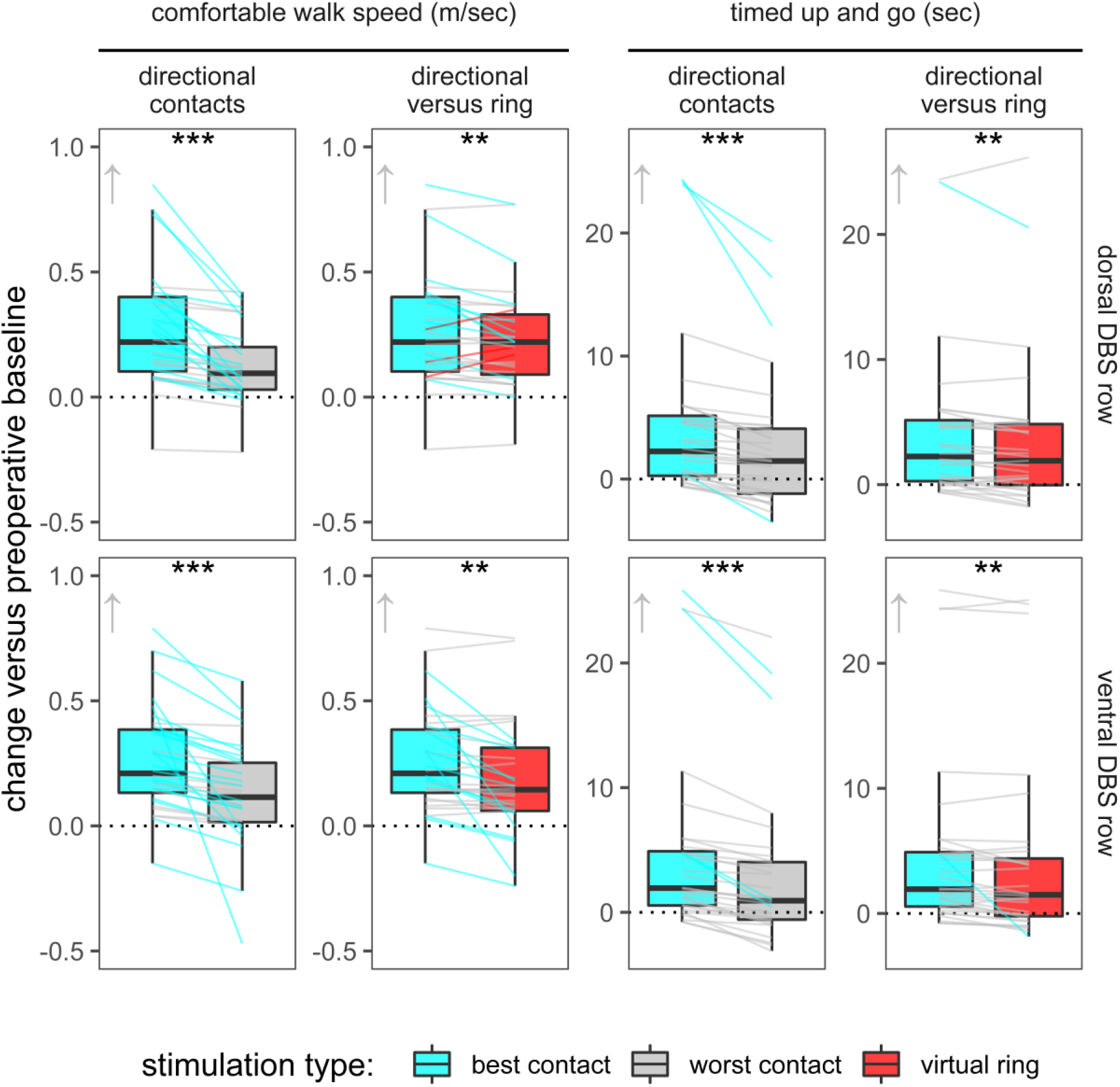
Comfortable walk speed and timed up and go measure gait function and overall mobility. Optimized directional stimulation yields more group-level motor improvement than ring stimulation versus preop baseline on both tasks (arrows indicate direction of improvement). Specific directional and ring stimulation parameters worsen gait and mobility in some individuals, and optimized directional DBS often rescues this worsening (line segments). Individual changes that meet or exceed the clinical important differences with directional versus ring stimulation are colored cyan and red, respectively. * p <0.05, ** p<0.01, and *** p<0.001 for group-level contrasts.

**Figure 3.**
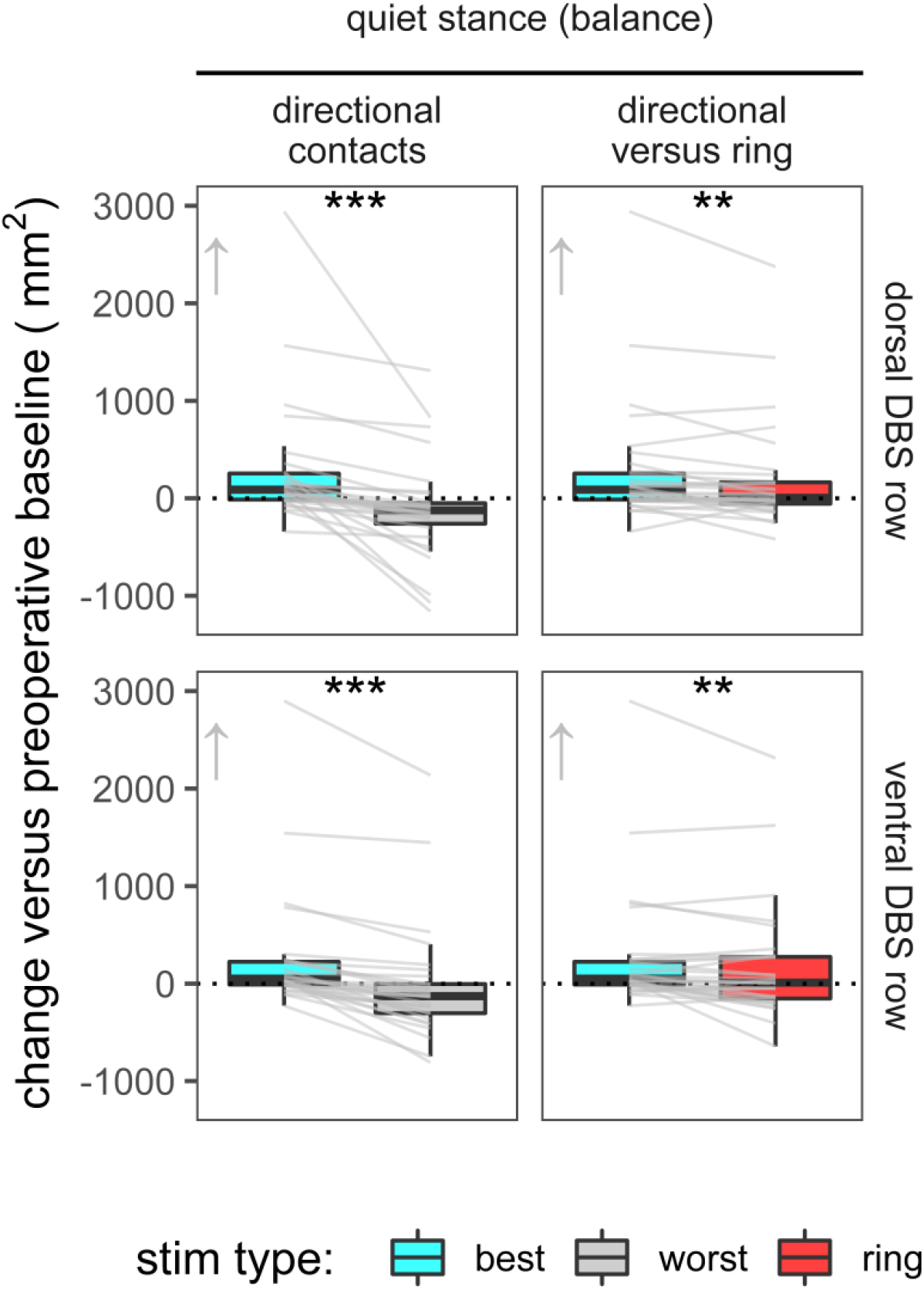
Static balance measured by an embedded force plate in the floor. Optimized directional stimulation yields greater improvement than ring stimulation on both tasks at the group level (boxplots). Despite overall improvements versus baseline, directional and ring stimulation can both worsen performance, and optimized directional DBS often rescues this worsening (line segments). Clinically important differences are not established for this task. * p <0.05, ** p<0.01, and *** p<0.001 for group-level contrasts.

**Figure 4.**
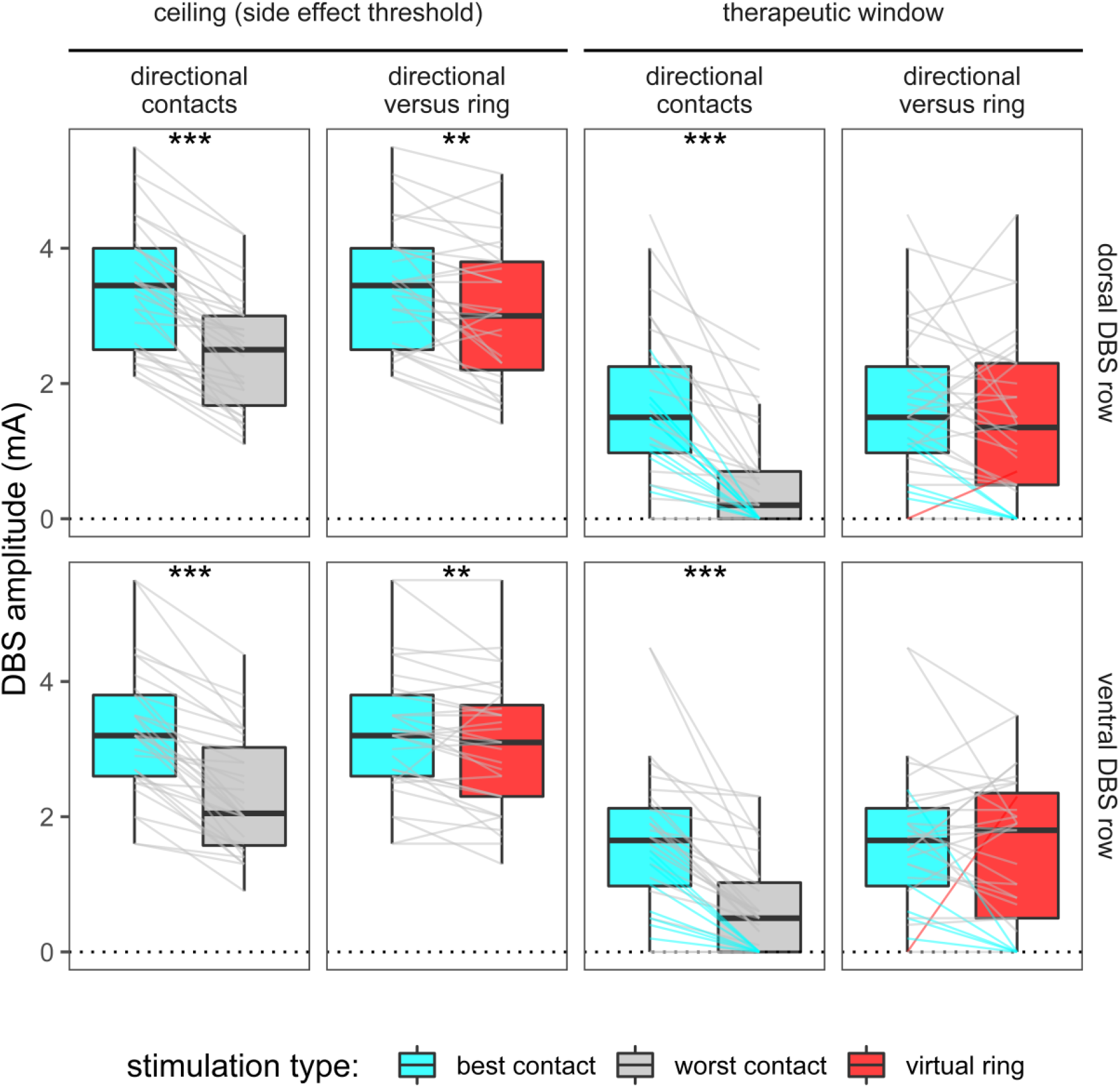
Directional stimulation improves amplitude thresholds for dose-limiting side effects and therapeutic window. Optimized directional DBS often rescues both virtual ring and directional configurations that display no therapeutic effects on the same row (cyan line segments). * p <0.05, ** p<0.01, and *** p<0.001 for group-level contrasts.

**Figure 5.**
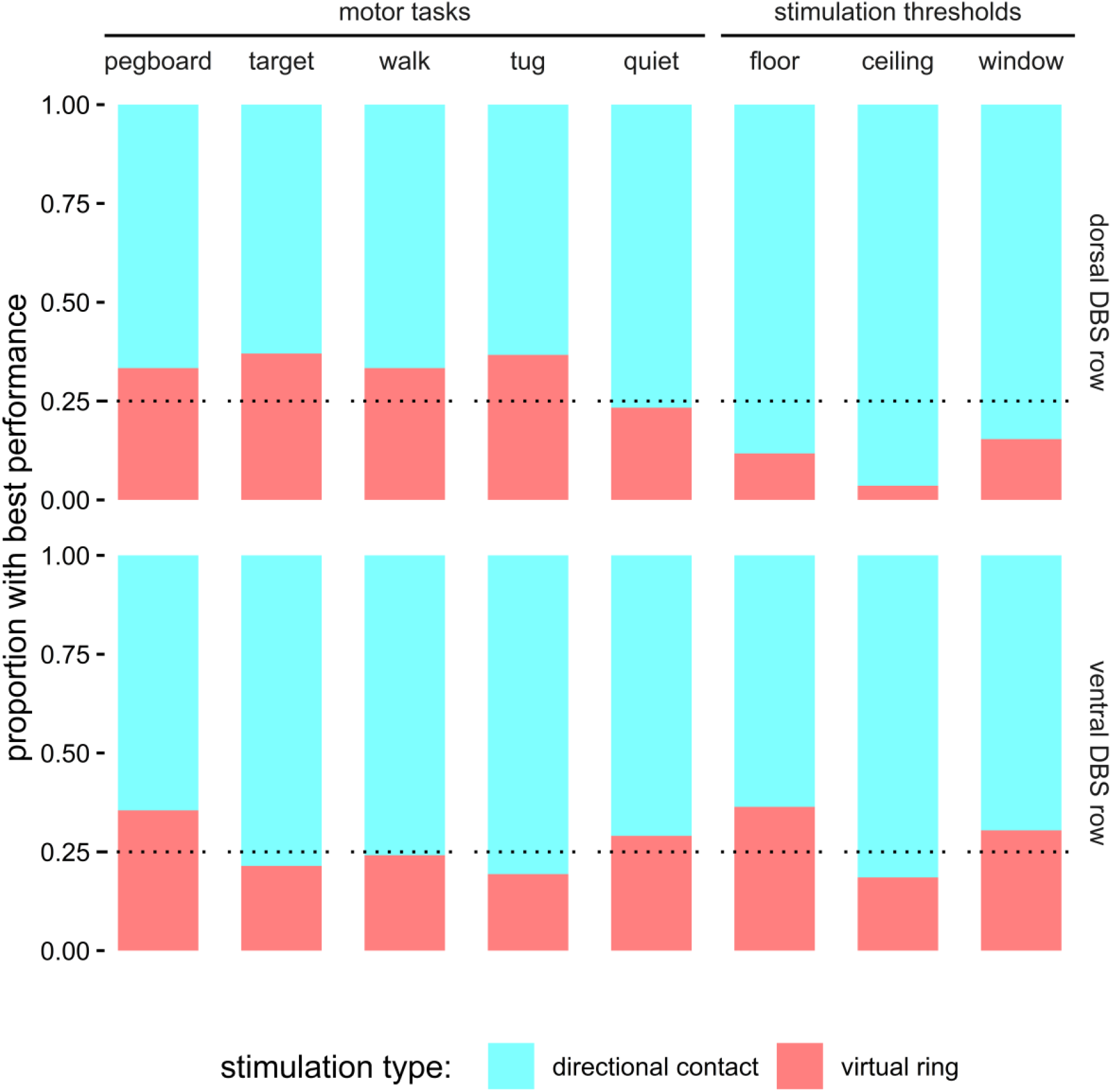
Optimized directional stimulation yielded the best behavioral outcomes more frequently than ring stimulation across multiple task demands. Dotted line represents chance within each directional DBS row (i.e., a given stimulation configuration would be selected among the three directional contacts plus their corresponding virtual ring within each DBS row at probability of 1/4 based on chance alone).

### Stimulation thresholds with directional versus ring stimulation

Stimulation thresholds (therapeutic window, floor, and ceiling) differ both among directional contacts and between the best directional contact and its corresponding virtual ring (Fig. 5). We measured significant main effects on side effect threshold but not therapeutic window on both DBS rows, whether across directional contacts or contrasting the best directional contact versus its virtual ring (*p*<0.01 and *p*=0.07). We again observed non-significant effects of row and row by contact interaction (*p*=0.59, *p*=0.28 and *p*=0.81, *p*=0.74) for side effect threshold and window, respectively).

### Predictors for motor improvement by directional stimulation

Given the added complexity and time demands of directional DBS programming, we sought to identify group-level predictors for changes in motor function (Fig. 6, Fig 7). We used R statistical packages ‘rmcorr’ to calculate repeated measures correlations across all directional contacts and tasks and ‘corrplot’ to generate matrix visualizations. The best predictor for motor function at a given directional contact was motor performance on the other tasks. Stimulation threshold items (floor, ceiling, therapeutic window) correlate strongly internally but predict motor performance only modestly. Broadband, low (13-20 Hz), and high (20-30 Hz) beta power from a given directional contact, whether rendered in common average reference or a variety of bipolar montages, did not predict therapeutic window or improvement on any of the motor tasks upon stimulation from that contact (Fig. 7).

**Figure 5.**
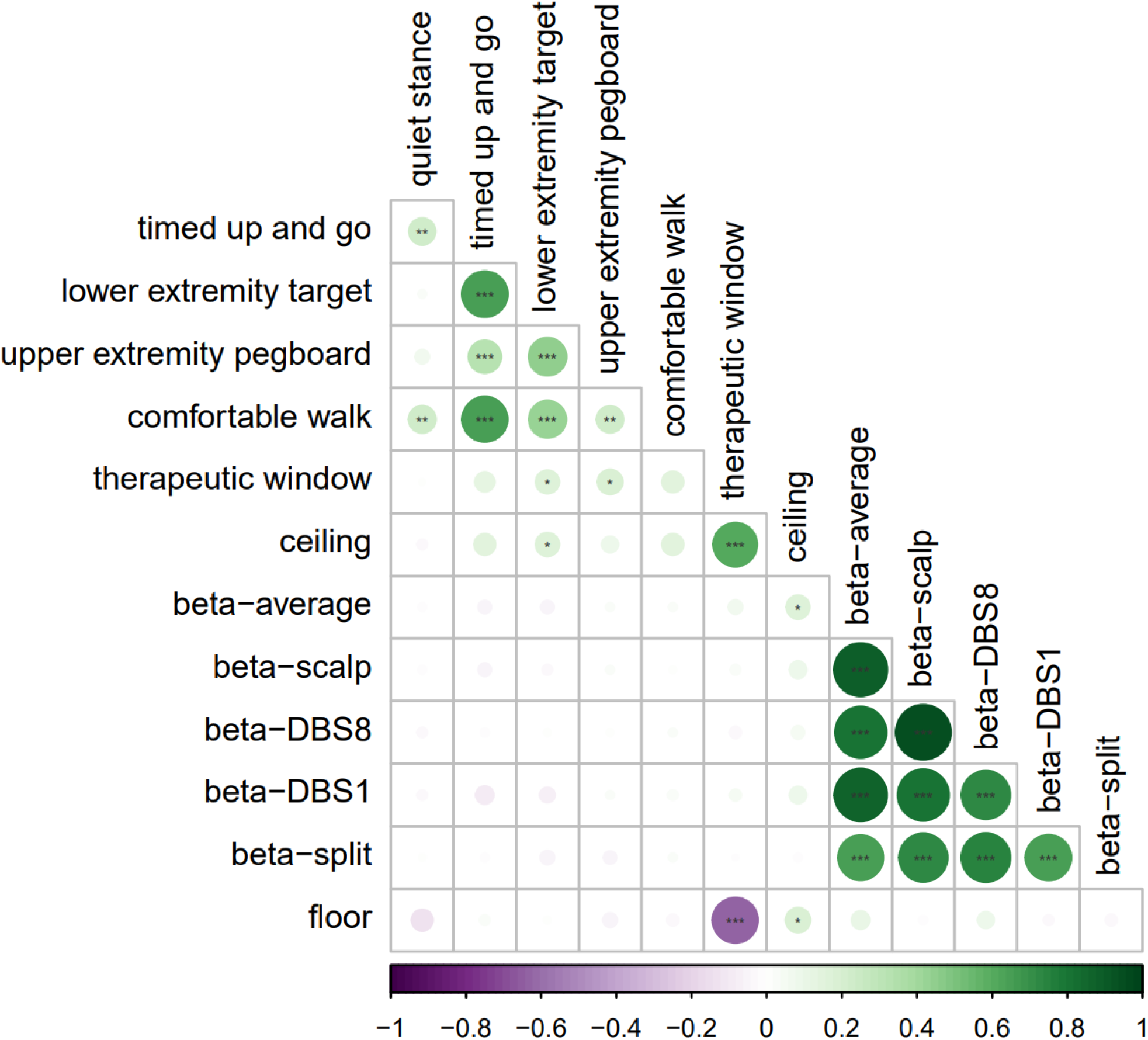
Matrix of repeated measures correlations of all dependent variables, organized in an unsupervised manner by ‘corrplot’ based upon their covariance structure. The best predictor for motor improvement from directional DBS is performance on the other motor tasks. Stimulation thresholds (floor, ceiling, and therapeutic window) correlate strongly internally but predict motor performance only modestly. Beta power referenced to common average, DBS contact 1, DBS contact 8, scalp, and split reference (dorsal and ventral contacts to DBS8 and DBS1, respectively) correlate strongly internally but do not predict motor improvements or stimulation thresholds. Color and dot size represent the correlation coefficients, and asterisks correspond to p-values (* p<0.05, ** p<0.01, and *** p<0.001)

**Figure 7.**
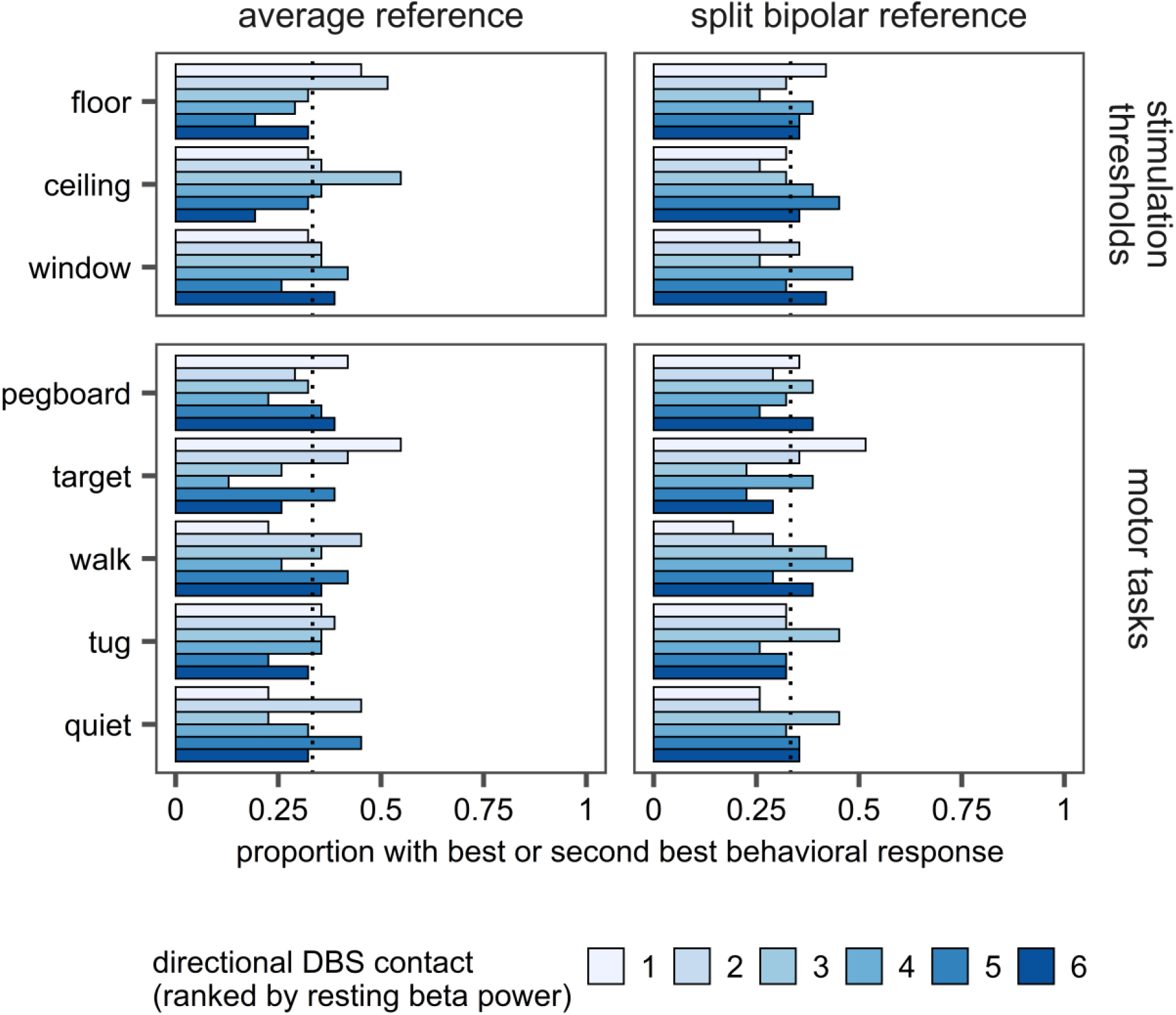
Resting beta power did not predict stimulation thresholds or motor improvements from directional DBS. Dotted line represents chance (i.e., a given rank would identify the best or second best behavioral outcome among the six directional contacts at probability of 1/3 based on chance alone). For these analyses, we rendered beta in both common average and split bipolar montage (dorsal and ventral contacts to DBS8 and DBS1, respectively).

## Discussion

Directional STN DBS elicits significant and in many cases clinically meaningful changes in quantitative, continuous measures of motor function during a double-blind randomized device activation, both within and across PD patients. In many instances, and across multiple functional domains, directional DBS improves or rescues suboptimal motor improvements from ring stimulation (Figs. 1-3). Further, the range of motor performance is typically larger for the worst versus best directional contact on a given DBS row. Collectively, our findings suggest that the spatial flexibility of directional leads not only improves tolerability but can likely yield better motor performance, as well.

Optimized directional stimulation improved motor function versus ring stimulation at the group level across each of five objective tasks evaluating upper and lower extremity dexterity, gait, mobility, and balance. These changes were measured in a randomized, double-blind manner with a first-generation directional lead, an essential step for better understanding its potential clinical impact. Prior literature on directional stimulation used one or a few UPDRS items (primarily contralateral arm rigidity) to define motor efficacy.^3,5,25^ Other studies measuring efficacy with ring-shaped DBS electrodes using continuous motor outcomes found effect sizes ranging from small or equivocal^26-29^ to large and clinically significant.^9,22^ While some elements of response variability might relate to lack of directionality with prior generation leads, other factors such as disease phenotype, electrode location, and predominant focus on upper limb function likely contribute as well.

Our findings confirm and expand on prior work suggesting that directional stimulation often improves amplitude thresholds for DBS side effects. Among the three directional contacts on a given row, side effect thresholds on a per-contact basis typically improve, are relatively unchanged, or worsen versus their corresponding virtual ring. Of note, some directional (and ring) contacts display no therapeutic window and/or worsened functional performance on quantitative motor tasks. Although their tissue activation volumes differ, the rationale for delivering ‘virtual ring’ stimulation when one or more of its directional contacts serves little or no therapeutic role in isolation is unclear. Also, instances of worsened motor function from directional DBS versus baseline were not an obvious outcome, given that we performed motor testing in the center of the therapeutic window. Thus, while reduction in rest tremor and/or rigidity are valuable, these findings might not always translate into improved functional performance with more complex real-world task demands. Regardless, novel algorithm-, neuroimaging-, or biomarker-based methods to guide directional programming should be validated with rigorous, systematic motor testing and implemented clinically with some caution, as directional stimulation can sometimes worsen motor performance.

Although DBS generally improves UPDRS motor items and more integrated functional tasks,^6,8,9,30^ outcomes can vary.^9,26^ Some of the observed variance might relate to not explicitly optimizing stimulation parameters based on motor performance. Directional stimulation provides opportunities to measure changes in performance in greater detail and across more diverse functional domains. Of note, directional DBS generally improves limb dexterity, gait, and overall mobility across participants, but balance (quiet stance) tended to worsen more readily than the other tasks. Various authors have proposed distinct functional sub-regions in the subthalamic nucleus,^31,32^ and STN stimulation does not improve balance measured in the lab or in the community as consistently as it improves gait and overall mobility.^28,33^ Our results suggest potential opportunities to exploit this topography, whether by enhancing efficacy or avoiding unintended and sometimes subtle declines in function.

How then do we harness potential advantages of directional stimulation? One challenge is that directional leads greatly enlarge the potential solution space for therapy within a given patient increasing the time to systematically optimize outcomes. This highlights the need for robust predictive biomarkers to guide DBS therapy. In our hands, the best predictor for motor efficacy at a given stimulation site was performance on another motor task (Fig 5). While sensible and internally consistent, this does not address the unmet need for dimension reduction in DBS programming. Stimulation thresholds (therapeutic window) predicted efficacy only modestly, whereas resting beta power rendered in myriad ways did not predict performance on any of the motor tasks. One difference between our study and prior efforts by other groups is that we defined efficacy based on functional performance on quantitative motor tasks rather than elements of the UPDRS.^34^ We utilized these validated, timed motor performance tasks in lieu of 10 serial repetitions of the UPDRS part 3 upper extremity rigidity item, anticipating that this would provide more diverse task demands, greater objectivity, and better sensitivity to change.

Our findings regarding resting beta power warrant further discussion, as well.^5,12^ One caveat is that we measured the field potentials during surgery, such that the signals might be perturbed by ‘microlesion’ effects. However, other groups found modest correlations between directional beta power and efficacy using similar intraoperative signals.^5,25^ Another consideration is potential lead rotation following surgery, however this is unlikely,^35,36^ as any rotations are typically considerably less than the angular dimensions of a single directional contact. Given the mixed findings from various groups, correlations between local field potentials and motor efficacy should be studied prospectively in larger samples and with chronic recordings >4 weeks after implant. Importantly, even if beta cannot consistently inform selection of directional contacts, it is nevertheless an important component of field potential signals arising from central motor circuits^37,38^ and still might prove useful as a control signal for adaptive stimulation.^39^ Also, the asymmetric 1-3-3-1 design of first generation directional electrodes complicates interpretation because the surface area of the ring contacts (1 and 8) differs substantially from the smaller directional contact segments.^40^ Future lead designs with a more symmetric array of directional contacts should allow more balanced sampling to better inform contact selection, particularly in the vertical axis of the DBS lead. Alternative methods to guide DBS programming arising from novel candidate biomarkers,^41^ device innovations such as beam-searching (unavailable at the initiation of our study),^42^ and novel predictive algorithms^40^ warrant further investigation, as well.

Our study has some potential limitations. Repeated motor assessments might be associated with behavioral responses that linger across conditions; however in prior work, we found that comfortable walk speed created large differences in motor performance with high reliability across DBS contacts.^9^ Additionally, serial assessments of motor function are ecologically valid in this context, as they remain the gold standard for decision-making related to clinical DBS programming. Future studies should examine whether motor improvements are sustained over longer time intervals. Another consideration is that we only studied directionality with single contact segments. Finer gradations of current fractionation across multiple contact segments should be evaluated, as well. Finally, we only investigated PD patients who received DBS in the STN, which typically has less favorable side effect thresholds than other stereotactic targets.^43^ Directionality likely confers different advantages or disadvantages for other brain locations or disease states.

## Conclusions

Optimized directional STN DBS can yield better motor performance than conventional ring stimulation, in addition to known advantages related to tolerability. Robust predictive biomarkers are needed to efficiently identify optimized directional stimulation parameters.

## Data Availability

All data produced in the present study are available upon reasonable request to the authors

## Acknowledgments

Funding Dr. Walker receives research funding from the National Institutes of Health (1UH3NS100553-38401), the Michael J. Fox Foundation (MJFF 15098)

## Data Availability

All data are available on Data Archive for the Brain Initiative (https://dabi.loni.usc.edu/home)

## Author contributions

A. Experimental design.

B. Data collection.

C. Data analysis and/or creating figures.

D. Writing of manuscript.

E. Edits of manuscript.

Christopher P. Hurt: A, B, C, D, E

Daniel J. Kuhman: A, B, C, E

Joseph W. Olson: B, C, E

Arie Nakhmani: A, C, D, E

Melissa H. Wade: B,

Bart L. Guthrie: B, E

Harrison C. Walker: A, C, D, E

